# Impact of real-time assessment on the training of trainers for the introduction of rotavirus vaccine in India

**DOI:** 10.1101/2021.07.20.21260191

**Authors:** Amanjot Kaur, Arindam Ray, Seema Singh Koshal, Syed F Quadri, Mayank Shersiya, Pradeep Haldar, Sanjay Kapur, Mohammed Haseeb, Arup Deb Roy

## Abstract

**Objectives:** To assess the effectiveness of training workshops for knowledge enhancement of program managers prior to rotavirus vaccine (RVV) introduction in the routine immunization program.

**Method:** The study was conducted among the participants attending two training workshops for the introduction of RVV; a state workshop in Pune and a regional workshop in Guwahati. The participants who attended the workshops and participated in both the pre and post-test - 53 for Guwahati and 59 for Pune. Data was collected in real-time via pre-post-test.

**Results:** In both workshops, a comparison of pre-test and post-test scores of all questions taken together showed a significant increase in the knowledge level of the participants (p<0.05). In Guwahati, the knowledge of the participants regarding doses of RVV, inadequate dosing, Vaccine Vial Monitor (VVM), open vial policy, operationalization of RVV, and monetary incentive increased significantly. In Pune, the knowledge of the participants regarding doses of RVV, bundling approach, schedule and dose, storage temperature for Rotavirus vaccine, VVM, open vial policy, vaccine delivery, and operationalization of RVV increased significantly after the training.

**Conclusions:** A pre-planned and well-designed knowledge assessment tool can be used to understand the impact of training workshops in enhancing the knowledge and practical skills of the participants prior to the introduction of a new vaccine.

## Introduction

In March 2016, India introduced the rotavirus vaccine (RVV) sub-nationally to reduce the enormous disease burden due to rotavirus diarrhea in under-five children^1^. In 2019, RVV was scaled up to the remaining 26 states and union territories (UTs), covering the residual 44% of India’s birth cohort.

Despite the increasing demands placed on the public health workforce due to the introduction of several newer vaccines, skill deficits are evident and may reflect inadequate preparation leading to on-job trial and errors. Hence, an important pre-requisite of any new vaccine introduction is the capacity building of the health functionaries, from the program managers to the frontline health workers (FLWs). Studies have shown that the knowledge, attitudes, and practices of the health care providers, including the frontline workers and supervisors, are the determining factors for improving immunization coverage^2-7^.

However, limited research is available on the effectiveness of capacity-building interventions related to capacity building of health functionaries. To our knowledge, an evaluation of the effectiveness of capacity-building interventions in new vaccine introduction has not been done so far. Evidence related to the effectiveness of these strategies will go a long way in considering choices about strategies and training focus, leading to better training outcomes^6^.

The pre-test and the post-test pattern help in evaluating the enhancement in the knowledge of the participants. The pre-test has also demonstrated that it improved the focus of the audience in those technical sessions/topics, which they could not answer correctly during the pre-test^8^. It also enabled the facilitators to emphasize specific topics during the technical sessions based on the question-wise scores’ analysis of the pre-test.

Currently, there is limited evidence available from India and other regions of the world measuring the impact of training via pre-and post-assessment in the context of immunization and other public health programs. Also, there is a dearth of literature evaluating the efficacy of short-term training programs in new vaccine introductions.

Hence, to bridge this evidence gap, for the first time, the authors have attempted to assess the effectiveness of training in rotavirus vaccine introduction evaluating the knowledge enhancement of the participants. The study results will enable the public health professionals and program managers to develop tailor-made training modules in new vaccine introductions to ensure a better application of knowledge during the rolling out of new vaccines.

In the present study, the authors have evaluated the effectiveness of a regional and a state training workshop for the introduction of RVV through understanding the level of knowledge enhancement before and after the conclusion of the workshop.

## Methodology

### Study participants and sampling technique

This study was conducted amongst participants attending a state-level ToT in Pune and another group of participants attending a regional ToT in Guwahati to introduce RVV in their respective regions. The sampling technique used in the study was the complete enumeration method. Table 1 depicts the demography of the participants in the two ToTs included in the study.

**Table 1:**
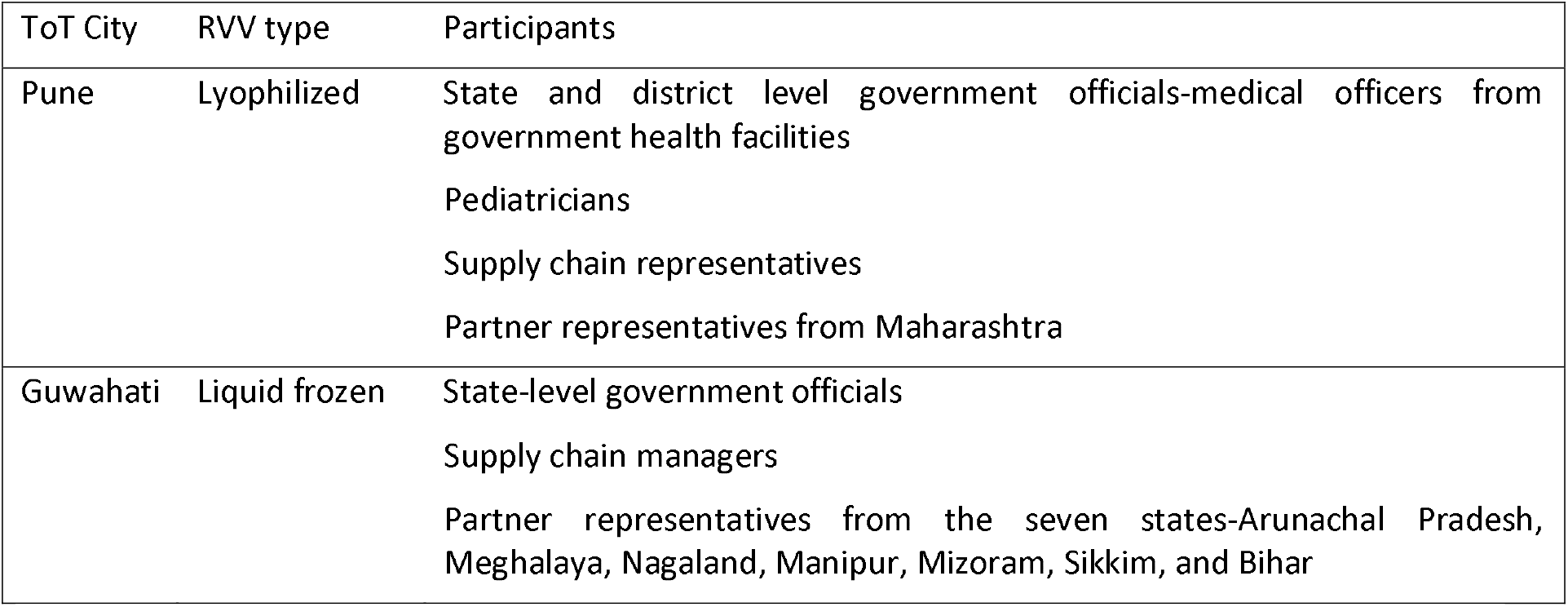
Demography of the participants at the two study ToTs (Pune and Guwahati) ToTs: Training of trainers

The Pune state ToT participants included state and district level government officials, including medical officers from government health facilities, pediatricians, supply chain managers, and partner representatives from Maharashtra.

In the regional ToT at Guwahati, the participants were only state-level government officials, supply chain managers, and partner representatives from the seven states, namely Arunachal Pradesh, Meghalaya, Nagaland, Manipur, Mizoram, Sikkim, and Bihar.

### Inclusion and exclusion criteria

All participants in the two workshops who participated in both the pre and post-test, respectively. The participants who participated either only in the pre-test or only in the post-test.

### Ethics approval

Ethics approval was obtained from the MGM-ECRHS, MGM Medical College, Aurangabad, Maharashtra (Ref.No.MGM-ECRHS/2019/16).

This study is an assessment of the pre and post scores of the participants attending a Government organized training workshop (supported by development partners). The government authorities nominated the participants, and the standard agenda was shared with them before the workshop. Also, before administering the pre and post-test, the participants were briefed by the facilitators during the workshop. The participants, by submitting their answers, gave their informed consent. The analysis of the proportion of question-wise correct responses was shared with the participants’ real-time.

### Training of trainers

In the year 2019, before the scale-up of RVV, a cascading approach was adapted to train health functionaries at all levels from regional through state and district to sub0district level. This model was conceptualized to complete the training of many healthcare workers in a short period of 2 months. To train the master trainers at the regional and state level, a national pool of facilitators comprising senior officials from the Ministry of Health and Family Welfare (MoHFW), immunization partner representatives, and senior faculties from academic or research institutions was formed. A total of seven training of trainers (ToTs) were conducted at different geographical zones, and a pool of 778 master trainers was created. Out of these seven ToTs, four were conducted to introduce lyophilized RVV at Pune, Hyderabad, Ahmedabad, and Kolkata, while three ToTs were held at Guwahati, Chandigarh, and Raipur for the introduction of liquid frozen RVV. The training modules were appropriately customized for the introduction of the two different of RVVs.

The master trainers trained at the regional/state level facilitated the state, district, and sub-district training. In these ToTs, the focus was on the capacity building of the participants. Each ToT was accompanied by assessing the participants’ knowledge both before and after the technical sessions through a pre-and post-test questionnaire.

### Study technique

The pre and post-test were conducted as per the standardized agenda approved by the MoHFW. The pre-test was conducted using a self-administered questionnaire before the start of the workshop to assess the participants’ knowledge of few program domains essential for introducing the RVV. The facilitators first briefed the participants on the objective and the process of conducting the pre-test. The questionnaire, which had ten questions, was then administered online using a google form. At the end of all technical sessions on day 2, the post-test using the same questionnaire was again self-administered to the participants on the google form.

Questions were close-ended with one correct answer for each question. The number of options for each question ranged from 2 to 5. The participants were allotted 20 minutes to complete the tests. To prevent data contamination, facilitators ensured that there was no discussion amongst the participants and that doubts, if any, were clarified individually. The participants were not allowed to refer to any training materials like operational guidelines for answering the questions. The participants had to answer every question before submission.

As the tool was linked to a live google form, so real-time analysis of the responses was done. In the final wrap-up session of the workshop, those questions where the proportion of correct responses were below 90% were again discussed, and the facilitators clarified all doubts of the participants.

To capture the difference in the formulation and the mode of administration of the two types of RVV, question numbers 2,3 and 4 were different between the questionnaires used in the Guwahati and Pune ToTs.

### Statistical analysis

As the google form also captured each participant’s name and mobile number to create the unique field, so for each workshop, a database of participant-wise responses in pre and post-test was available. To calculate the pre and post-test scores, correct and incorrect responses to each question were assigned scores of 1 and 0, respectively. The pre and post-test results were then plotted in a simple bar graph to compare the difference in the proportion of question-wise correct responses. Then, to understand whether these differences in correct responses in each question and the overall difference in correct responses were statistically significant, three tests of significance were done. These tests were administered separately on the data compiled for each of the two ToTs. Statistical package for the social sciences (SPSS) version 20.0 (Chicago, SPSS Inc) was used for the final analysis.

The number of correct responses of the participants before and after the workshop-captured through the pre and post-test, was compared for statistical significance using McNemar’s test. As the scores could not be assumed to be normally distributed, Wilcoxon Signed Rank Test was administered to test the significance of the change in the participants’ level of knowledge, as measured by the median total score. We also found several ties in scores between the pre and post-test, which may potentially dilute the comparison of the median score, as demanded by the Wilcoxon signed-rank test. Hence, paired t-test was applied to further validate the results.

## Results

The ToTs were 2-day workshops with a structured and standardized agenda and a customized adult learning-based training methodology. A total of 84 and 60 participants attended the ToTs at Pune and Guwahati, respectively. However, the total number of participants who completed both the tests was 59 and 53 participants in the Pune and Guwahati workshops.

### Regional ToT at Guwahati

In the regional ToT held in Guwahati, it was found that the knowledge about doses of RVV in one vial (77%), inadequate dosing (28%), VVM (68%), phasing in of RVV in the immunization schedule (74%) and full immunization incentive for ASHA (68%) were the least amongst the participants. Table 2 shows the difference in the participants’ level of knowledge before and after completing the ToT. It also shows whether the difference in the knowledge level is significant or not. The analysis of post-test data revealed that the proportion of correct responses increased to more than 90% in all the questions except that on inadequate dosing (66%).

**Table 2:**
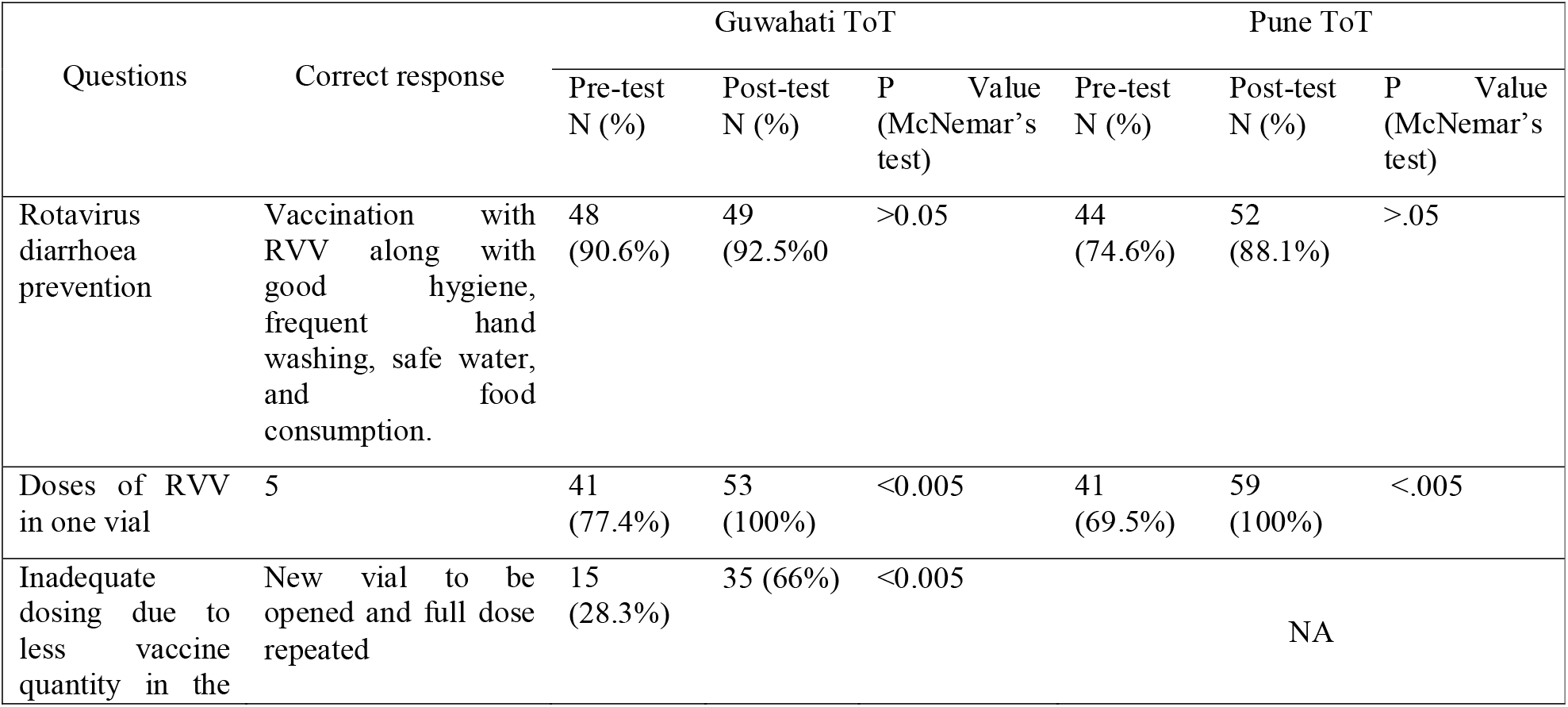
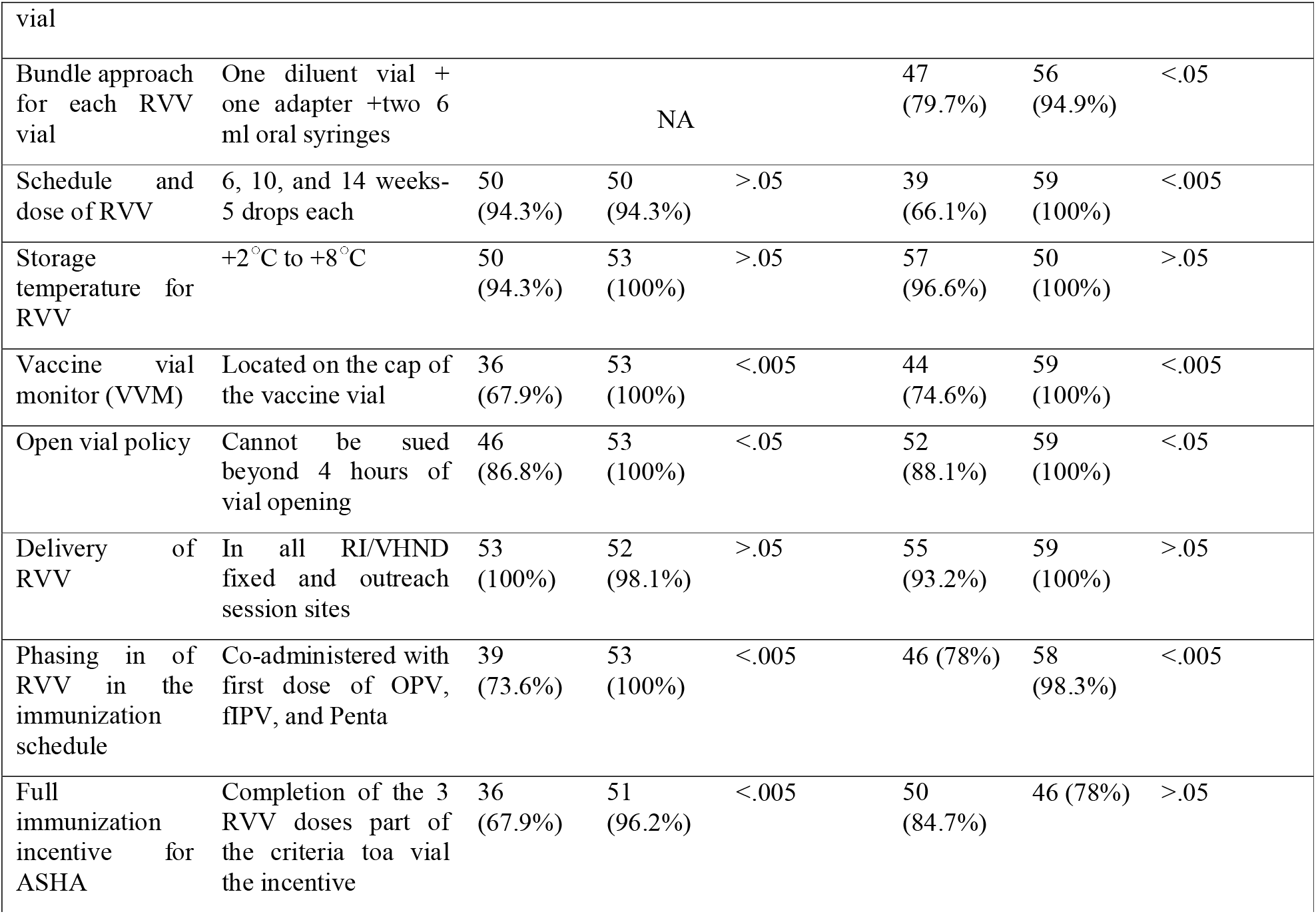
Findings from McNemar’s Test (Guwahati and Pune ToT) RVV: Rotavirus Vaccine; ASHA: Accredited Social Health Activist

The knowledge regarding doses of RVV in one vial (77.4%), inadequate dosing (28.3%), VVM (67.9%), open vial policy (86.8%), phasing in of RVV in the immunization schedule (73.6%) full immunization incentive for ASHA (67.9%) increased significantly to 100%, 66%, 100%, 100%, 100% and 96.2% respectively.

Table 3 shows the change in the overall level of knowledge before and after completing the training workshop. Wilcoxon Signed-Rank test revealed a statistically significant increase in knowledge following the completion of the workshop, Z=-5.779, p<0.05, with a large effect size (r=0.94). The median knowledge score increased from 8 to 10 after the training workshop.

**Table 3:**
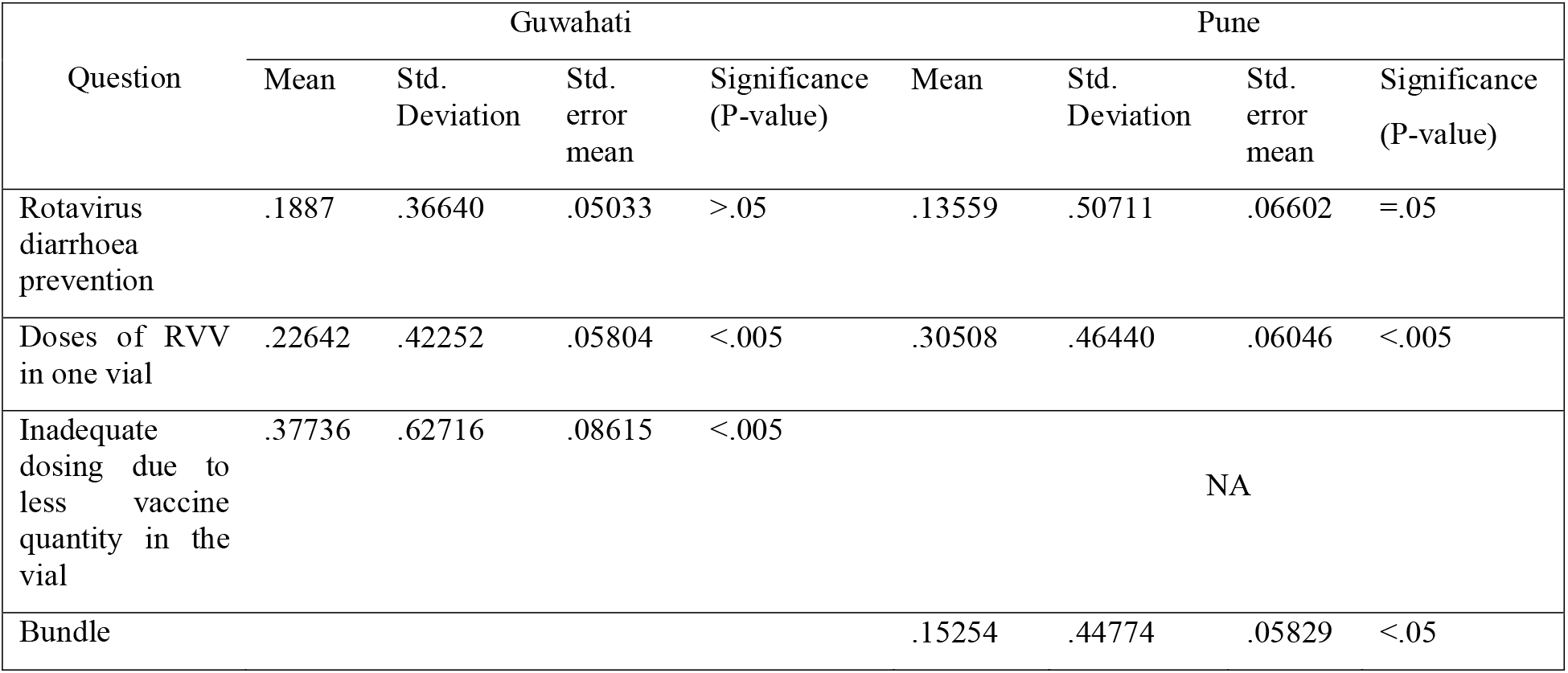
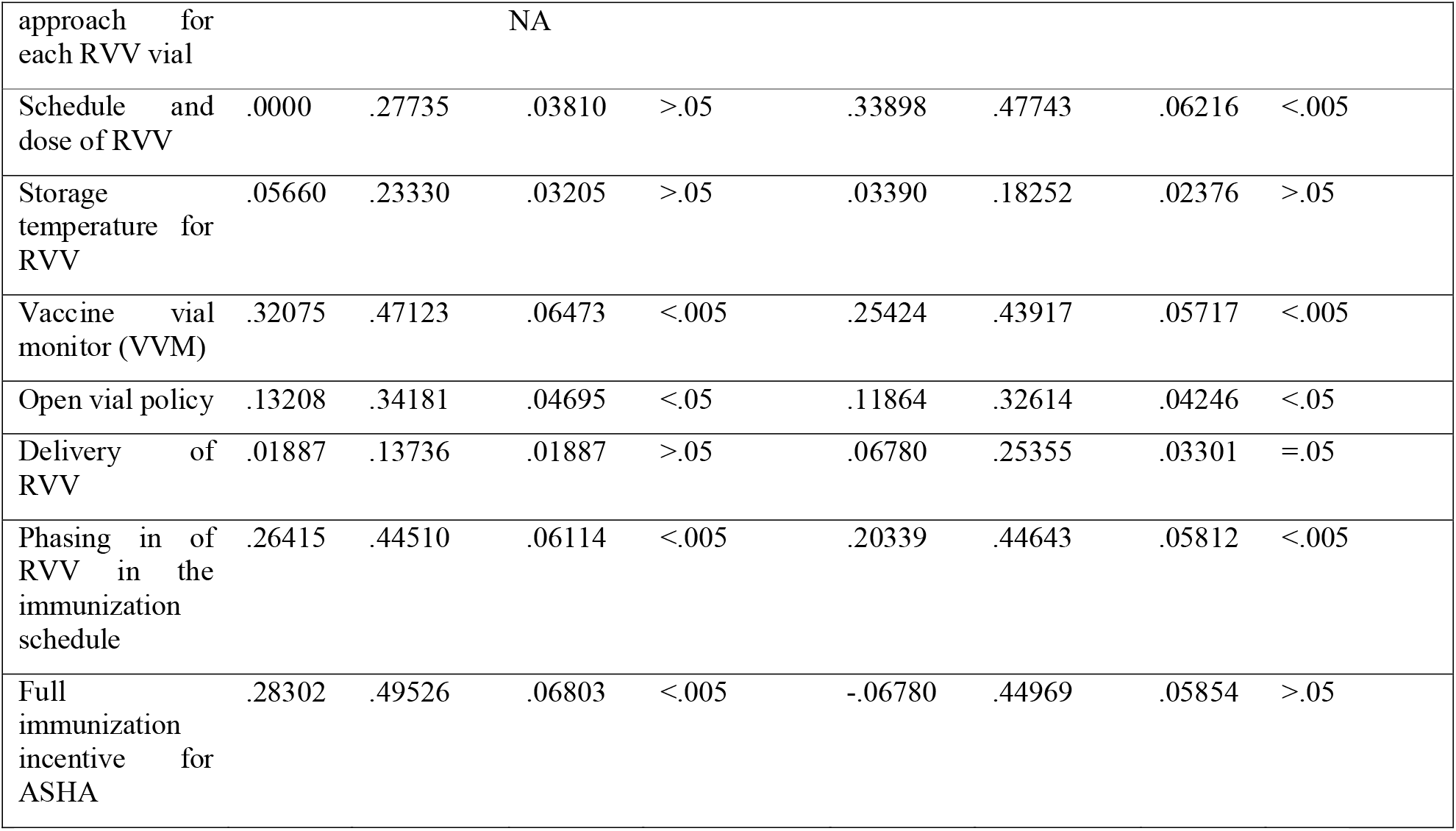
Findings from Paired T-test (Guwahati and Pune) RVV: Rotavirus Vaccine; ASHA: Accredited Social Health Activist

Table 4 shows the change in the level of knowledge for each question before and after completing the training workshop. Paired t-test revealed a statistically significant increase in knowledge following the completion of the workshop in knowledge around doses of RVV in one vial (p<0.005), inadequate dosing (p<0.005), VVM (p<0.005), open vial policy (p<0.05), phasing in of RVV (p<0.005) and full immunization incentive for ASHA (p<0.005).

**Table 4:** Wilcoxon Signed Rank Test (findings for Guwahati and Pune Tot)

| Score | Guwahati |  | Pune |  |
| --- | --- | --- | --- | --- |
|  | Pre-test | Post-test | Pre-test | Post test |
| Median | 8 | 10 | 9 | 10 |
| Interquartile range | 7-9 | 9-10 | 6-10 | 9-10 |
| Wilcoxon test (Z) | -5.779 |  | -4.996 |  |
| P value | <.05 |  | <.05 |  |
| Correlation coefficient | 0.94 |  | .73435118 |  |

### State ToT at Pune

In the state ToT held in Pune, it was found that the knowledge about schedule and dose of RVV (66.1%), doses of RVV in one vial (69.5%), rotavirus diarrhea prevention (74.6%), and Vaccine Vial Monitor (74.6%) were the least amongst the participants. Table 2 shows whether, for each of the questions, the difference in the level of knowledge of the participants before and after the completion of the ToT was significant or not. After the completion of all technical sessions in the workshop, the data captured in the post-test revealed that the proportion of correct responses increased to more than 90% in all the questions except the ones on rotavirus diarrhea prevention (88.1%) and full immunization incentive for AHSA (78%).

The knowledge regarding doses of RVV in one vial (69.5%), bundle approach for each RVV vial (79.7%), schedule and dose of RV (66.1%), storage temperature for RVV (96.6%), Vaccine Vial Monitor (74.6%), open vial policy (88.1%), delivery of RVV (93.2%) and phasing-in of RVV in the immunization schedule (78.0%) increased significantly to 100%, 94.9%, 100%, 100%, 100%, 100%, 100% and 98.3% respectively.

Table 3 shows the change in the overall level of knowledge before and after completing the training workshop. Wilcoxon Signed-Rank test revealed a statistically significant increase in knowledge following the completion of the workshop, Z=-4.996, p<0.05, with a large effect size (r=0.73). The median knowledge score increased from 9 to 10 after the training workshop.

Table 4 shows the change in the level of knowledge for each question before and after completing the training workshop. Paired t-test revealed a statistically significant increase in knowledge following the completion of the workshop in knowledge around doses of RVV in one vial (p<0.005), schedule and dose of RVV (p<0.005), VVM (p<0.005), and open vial policy (p<0.05).

## Discussion

This study was conducted to assess the effectiveness of ToTs in increasing the awareness and practical knowledge of the participants. A comparison of the pre-test and post-test scores on all questions together showed an increase in correct responses from 78% to 95% in the Guwahati workshop and an average increase from 81% to 96% in the Pune workshop. The post-test average correct responses were similar in both the workshops, although the participant profile was different-state-level program managers in Guwahati and sub-state or district level program managers in the Pune workshop. The increase in the knowledge level in both workshops is statistically significant (p<0.05). Uskun et al., in their study, concluded that training on immunization increased the knowledge of primary healthcare workers and the vaccination coverage in the study region. THE results of the study showed that the technical sessions during the training led to a significant increase (p<0.01) in the health workers’ knowledge of immunization. It was also seen that the vaccination coverage increased significantly (p<0.001) over three months post the training-based intervention^9^. There is adequate evidence establishing the positive effects of a training intervention that reflects an increased score of the post-test compared to the pre-test^10,11^.

While conducting a capacity-building workshop helps in familiarizing the participants with the various aspects and domains of the new vaccine introduction, checking the effectiveness of the technical sessions in enhancing the actual knowledge of the participants is an important activity during the workshop. There is a dearth of studies to assess the impact of training the cadre of supervisors or future master trainers. World Health Organization (WHO) conducted a study to evaluate the training system and the processes followed in some good and bad performing states in terms of immunization rates. In the study by WHO, Das et al. suggested that the quality of the training in terms of large pools of facilitators, prior knowledge, training methodology, and trainees’ feedback were found to be salient enabling factors in such trainings^12^.

The overall study emphasizes the importance of a knowledge assessment format for the training in public health. On the one hand, the pre-test individually is important for both the participants and the facilitators of the training; (i) to intimate the participants towards the various topics to be covered during the training, (ii) to highlight in their minds those topics beforehand in which they scored less in the pre-test; (iii) for facilitators to adopt a more interactive and easier approach toward the same topics where the scores were less during the pre-test. The post-test, on the other hand, individually imparts confidence among the participants when they score well. Together, the pre-test and post-test, when viewed comparatively, gave the actual picture about the quality of the training.

An advantage of this study is that it methodologically deployed an ensemble approach by applying the three most appropriate tests of significance in a hierarchical way to assess the trainees’ actual difference between pre and post training knowledge status. Though each method suffers from its limitations, this ensemble approach gives more confidence to the findings.

One limitation of this study is that it is a complete enumeration of the participants attending a particular workshop, and hence it may lack external generalizability. As the tests were done for “on the spot,” immediate training needs assessment and post-session feedback & clarification, our study also could not capture temporal change in participants’ knowledge. Though the results of this study show that following the intervention of a two-day training, a significant improvement is seen in the knowledge of the participants while answering the post-test as compared to the pre-test. However, the retention of this knowledge gain is not assessed after a long gap such as six months or one year. Similar studies have shown a progressive decline in the mean scores of the participants after three or six months of the intervention. Though a decline in knowledge is seen after the said time, the then scores were still higher than those before the training intervention^13,14^. In contrast to the findings mentioned above of declining knowledge, some studies have shown knowledge and skill retention after training^15^.

To create data-based evidence of the results of a training intervention, a periodic and systematic follow-up knowledge assessment after three months^16^, six months^17^, nine months^18^, 12, and 24 months^19^ should be included in planning such interventions. Also, it has been seen that for short-term results, a single intervention of continuing education can be useful, but for long-term sustainability and effectiveness, additional interventions to address health system gaps and community issues should be planned^20^.

## Conclusion

This study signifies and highlights the effectiveness of a pre-planned and well-designed knowledge assessment tool before and after a training workshop in bridging the knowledge gap and improving the practical approach needed for new vaccine introduction. The findings signify that the pre-test and post-test tool can be effectively used to capture the participants’ pre-training knowledge levels, suggest the key areas to focus during the training, and finally test the knowledge enhancement post-training. The post-test, being the last event of training, allows reinforcing the key take-home points. This study recommends that the same model of knowledge assessment through live pre-test and post-test approach should be used in all the training of Public Health. It will help make an accurate real-time assessment of the participant’s knowledge and make real-time corrections or modifications in the focus of the training to ensure the best outcomes in terms of knowledge and practical skills gained.

## Supporting information

Ethical Approval Page 1

Ethical Approval page 2

## Data Availability

All raw data can be made available whenever requested

https://www.example.com

