## Supplementary figures and images for "Impact of real-time assessment on the training of trainers for the introduction of rotavirus vaccine in India"

### Ethical Approval Page 1

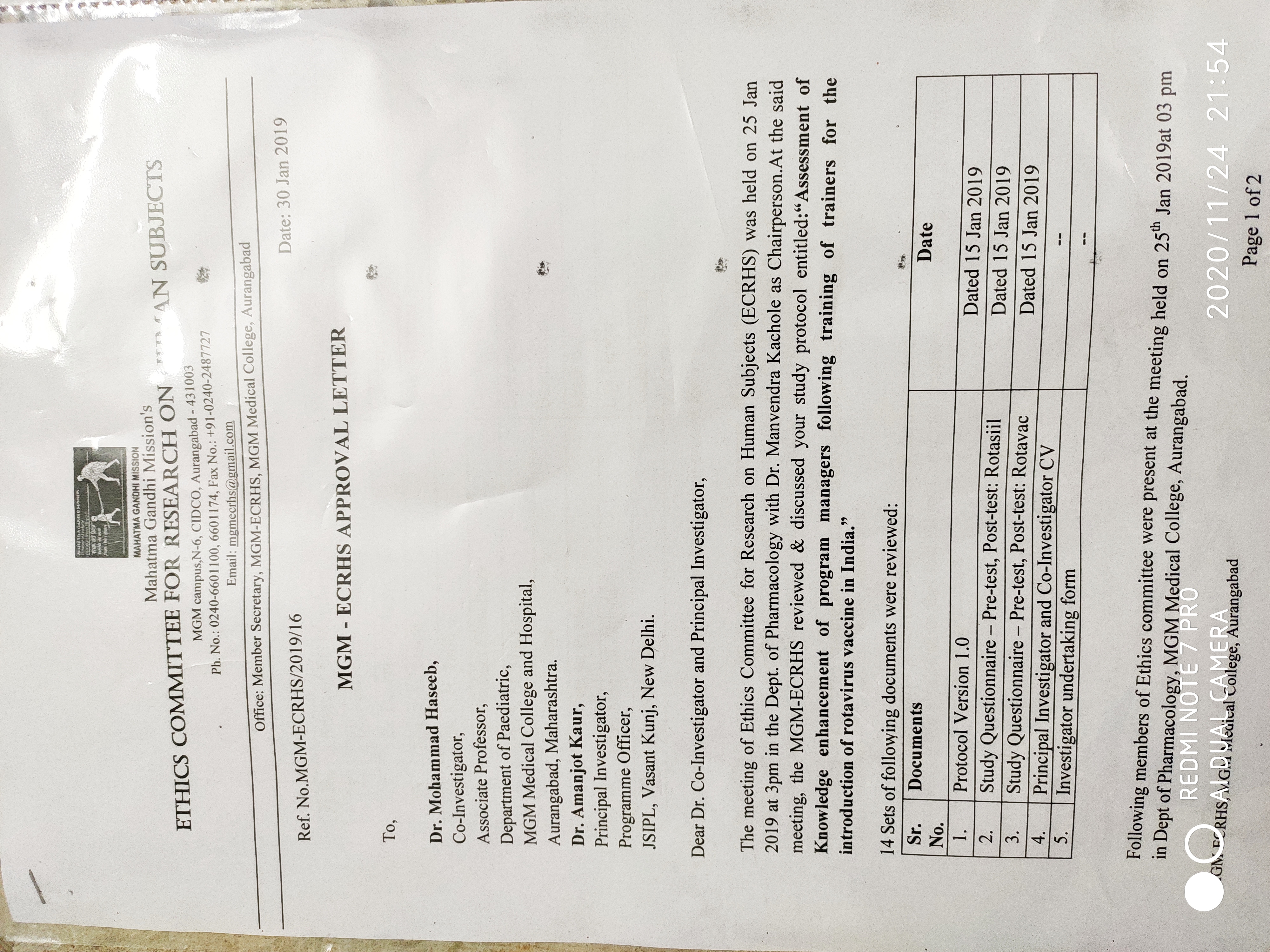

### Ethical Approval page 2

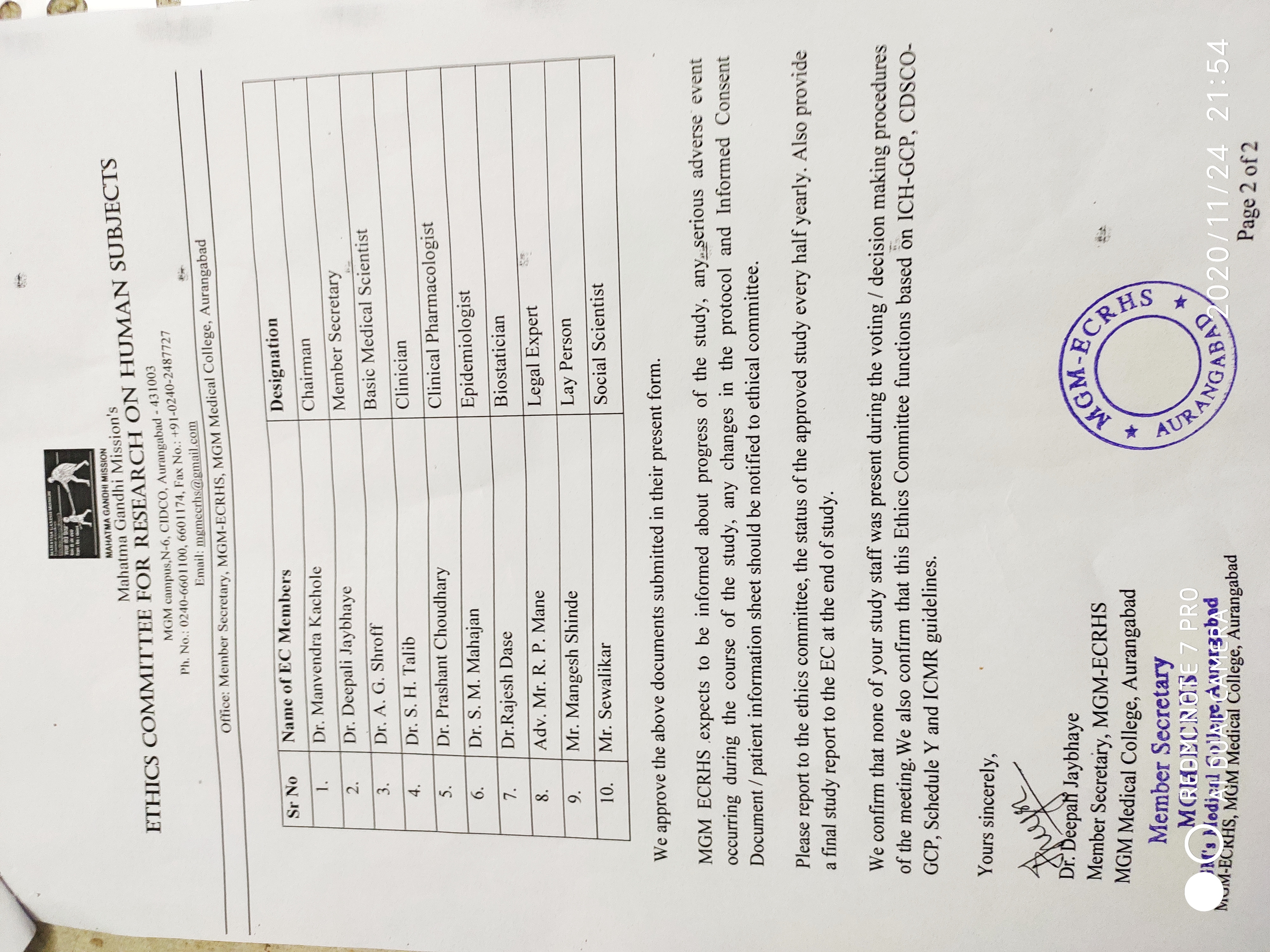
